# Reducing travel-related SARS-CoV-2 transmission with layered mitigation measures: Symptom monitoring, quarantine, and testing

**DOI:** 10.1101/2020.11.23.20237412

**Authors:** Michael A. Johansson, Hannah Wolford, Prabasaj Paul, Pamela S. Diaz, Tai-Ho Chen, Clive M. Brown, Martin S. Cetron, Francisco Alvarado-Ramy

## Abstract

Balancing the control of SARS-CoV-2 transmission with the resumption of travel is a global priority. Current recommendations include mitigation measures before, during, and after travel. Pre- and post-travel strategies including symptom monitoring, testing, and quarantine can be combined in multiple ways considering different trade-offs in feasibility, adherence, effectiveness, cost and adverse consequences. Here we use a mathematical model to analyze the expected effectiveness of symptom monitoring, testing, and quarantine under different estimates of the infectious period, test-positivity relative to time of infection, and test sensitivity to reduce the risk of transmission from infected travelers during and after travel. If infection occurs 0-7 days prior to travel, immediate isolation following symptom onset prior to or during travel reduces risk of transmission while traveling by 26-30%. Pre-departure testing can further reduce risk if testing is close to the time of departure. For example, testing on the day of departure can reduce risk while traveling by 37-61%. For transmission risk after travel with infection time up to 7 days prior to arrival at the destination, isolation based on symptom monitoring reduced introduction risk at the destination by 42-56%. A 14-day quarantine after arrival, without symptom monitoring or testing, can reduce risk by 97-100% on its own. However, a shorter quarantine of 7 days combined with symptom monitoring and a test on day 3-4 after arrival is also effective (95-99%) at reducing introduction risk and is less burdensome, which may improve adherence. To reduce the risk of introduction without quarantine, optimal test timing after arrival is close to the time of arrival; with effective quarantine after arrival, testing a few days later optimizes sensitivity to detect those infected immediately before or while traveling. These measures can complement recommendations such as social distancing, using masks, and hand hygiene, to further reduce risk during and after travel.

## Introduction

Coronavirus disease 2019 (COVID-19) was first recognized in late December 2019. By March 2020, the virus causing COVID-19, SARS-CoV-2, had reached 6 continents and almost 70 countries. In response to the global COVID-19 outbreak, governments implemented a variety of mitigation measures including unprecedented social distancing measures, travel health alerts, and travel restrictions at national and sub-national levels [1,2]. These measures, as well as concern about exposures related to travel, led to major and prolonged reductions in air travel worldwide [3–7]. Spatiotemporally asynchronous waves of COVID-19 have led to dynamic risk and mitigation measures globally with an accompanying interest in identifying risk management steps for travel that can reduce the risk of transmission and address concerns of travelers, travel industry regulators, and public health authorities [8–10].

Initial policies for managing translocation of the virus from one destination to another relied on closing borders or restricting entry of travelers from countries with higher incidence rates [11,12]. Although these approaches may have reduced the importation of some cases and preserved resources, they came with enormous economic and individual impacts [13,14].

For travelers, personal mitigation actions include wearing masks, social distancing at least 6 feet from others when possible, frequent hand washing or use of alcohol-based hand sanitizer, not touching their their face, and avoiding anyone who is sick. Governments, airlines, airports, and other businesses serving travelers have implemented or recommended measures to reduce the risk of COVID-19 associated with air travel [15,16]. These measures have included enhanced disinfection procedures, employee health assessments, passenger health attestations, screening for fever, illness response protocols and other steps to reduce risk of transmission in airports and on conveyances [10,17]. Symptom-based screening at airports has proven ineffective because those measures miss mild, afebrile, asymptomatic, and pre-symptomatic SARS-CoV-2 infections [18–21]. Asymptomatic persons may account for 20% to 40% of SARS-CoV-2 infections and can transmit the virus to others [22–26], and epidemiological data indicate that infectiousness begins prior to symptom onset for those who do develop symptoms [27–31].

In many destinations, arriving travelers, most of whom are asymptomatic with no specific known exposures, were asked to self-quarantine and reduce contacts as much as possible after arrival. The World Health Organization (WHO) defines quarantine as “the restriction of activities and/or separation from others of the suspect persons… who are not ill, in such a manner as to prevent the possible spread of infection” and indicates that quarantine may be considered for travelers based on risk assessment and local conditions. For known SARS-CoV-2 exposures, WHO recommends quarantine of 14 days based on the limit of the estimated incubation period for SARS-CoV-2 [32]. A 14-day quarantine alone, when effectively implemented and strictly adhered to, approaches 100% reduction in risk of transmission post-exposure [33,34]. However, travelers may have little incentive to consistently adhere to these measures at their destinations unless there is the ability to reliably communicate with them, support their needs, and enforce these measures. Monitoring and enforcing adherence to quarantine measures requires tremendous effort and resources by public health entities that may only be feasible in certain contexts.

Inclusion of SARS-CoV-2 testing as a component of a multi-layered approach to risk-reduction is currently being implemented in various settings. Some businesses and educational institutions are incorporating SARS-CoV-2 screening strategies into their concepts of operations, sometimes including mandatory testing of employees and voluntary testing for customers [35–37]. While there is no current international standard for testing travelers, many countries and jurisdictions are requiring arriving travelers to be tested either prior to their departure or after arrival to identify infected persons who are asymptomatic so they can be isolated [38]. Current guidance or requirements vary from country to country, and from state to state within the United States, including the timing of the test prior to or after travel, the type of test used (viral antigen, viral RNA), and the use of negative test results to alleviate additional public health measures, such as quarantine, at the destination [39,40].

Currently available SARS-CoV-2 tests for detecting active infections include nucleic acid amplification tests (NAAT), such as reverse transcription polymerase chain reaction (RT-PCR) tests, rapid isothermal NAATs (e.g., ID NOW(tm)), and rapid antigen-based tests. Time to deliver results is hours to days for RT-PCR and minutes to hours for rapid tests, which can also be processed without a specialized laboratory. Rapid antigen tests for SARS-CoV-2 are currently authorized in the United States for suspected cases in the first 5-7 days of symptoms [41,42]. While rapid antigen tests have advantages over NAATs in terms of cost, simplicity and turn-around time, there are limited data on their efficacy in screening asymptomatic individuals.

SARS-CoV-2 transmission risk related to travel can be viewed in two domains: transmission risk during travel (e.g., by infected travelers while at an airport or on aircraft) and after travel is completed (e.g., introduction of SARS-CoV-2 to the destination location). There is also overlap as transmission risk during travel can lead to new infections, which can increase post-travel risk. Data on strategies for reducing risk associated with travel are scant and there are many potential strategies (e.g., the optimal timing of pre-departure or post-arrival testing or the combination of testing and post-arrival quarantine) [36,43–45]. Mathematical models have provided some insights to the potential impact of quarantine combined with testing [44,46]. Here, we build upon those models, considering uncertainty in infectious periods and different testing options to assess a suite of possible combined pre- and post-travel strategies to reduce transmission risk from infected travelers.

## Methods

First, we characterized component processes related to transmission risk during infection: the relative infectiousness over the course of infection, the proportion of infections resulting in symptoms, the timing of symptom onset for those who have symptoms, and the probability of testing positive over the course of infection.

We used three distinct models to characterize relative infectiousness over time (Fig. 1a). We used a Gamma density function to approximate a 10-day infectious period with peak infectiousness on day 5 based on observations from numerous studies [47–51]. We also replicated a within-host infection model by Goyal et al. [52], which suggests that most people are infectious from days 3 to 7, with tapering afterwards. The final model characterizes simulated infectious periods from Clifford et al. [44], based on estimated latent periods (the delay between infection and becoming infectious) [47] and infectious periods [53]. The Goyal et al. and Clifford et al. infectiousness models were fitted by first simulating daily individual-level infectiousness profiles from the original code and then fitting density functions to the simulation data.

**Figure 1a.**
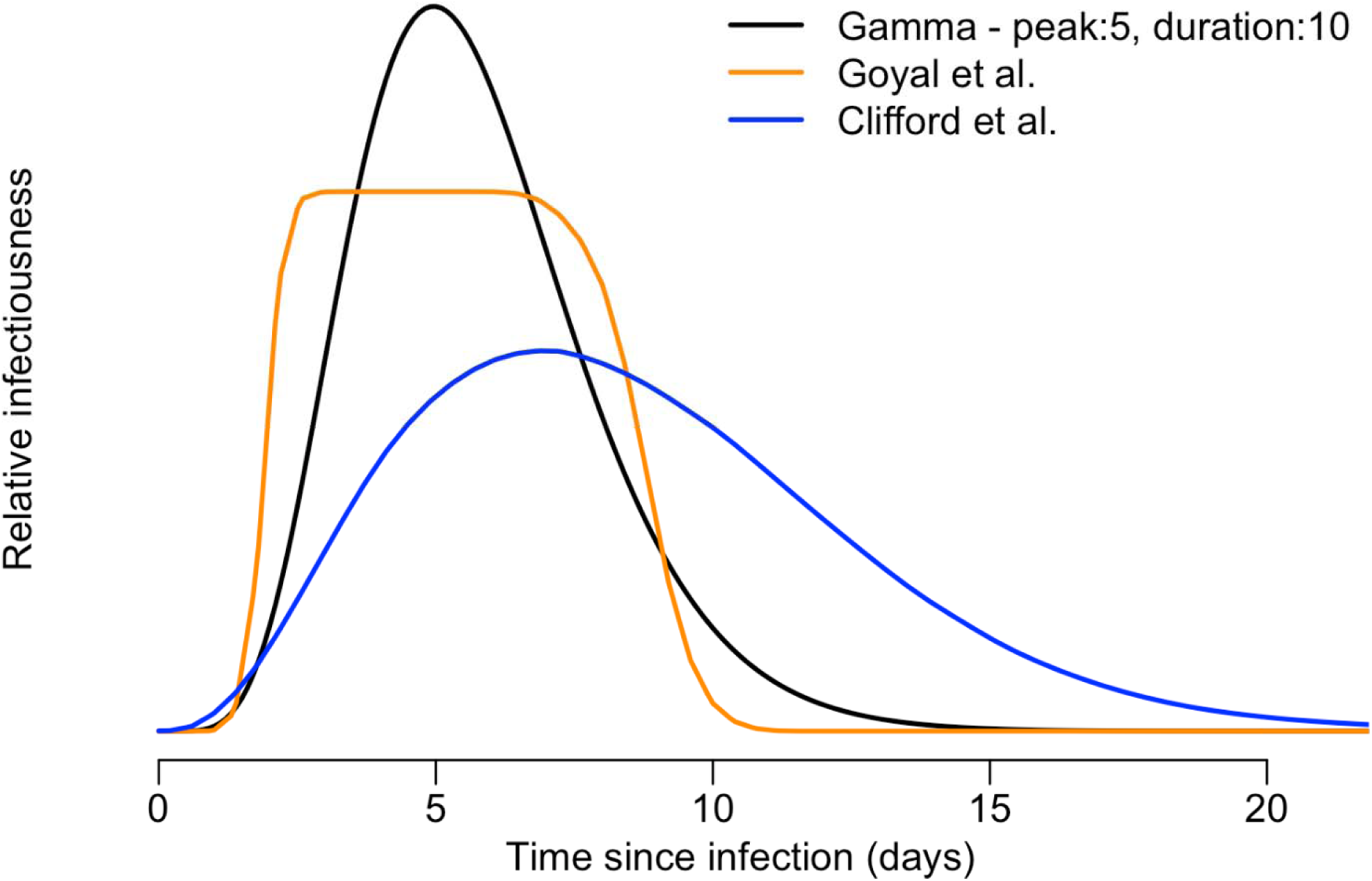
Models of average infectiousness of individuals infected with SARS-CoV-2 relative to time since infection: a Gamma density function approximating a 10-day infectious period with a peak on day 5 [47–51], a host infection model adopted from Goyal et al. [52], and simulated infectious and latent periods adopted from Clifford et al. [44].

We assumed that approximately 70% of all infections result in symptomatic COVID-19 cases [25]. For the incubation period, we used a meta-estimated median of 5 days with a Log-Normal distribution based on a meta-analysis by McAloon et al. [54].

For diagnostic testing, we used two models: one directly estimating positivity by RT-PCR and one approximating an antigen detection assay (Fig. 1b). For the RT-PCR model, we used the model generated by Clifford et al. [44] based on data from Kucirka et al. [55]. To approximate an antigen detection assay, we used the Gamma distribution for infectiousness as formulated above and calibrated it to have peak sensitivity of 80% at peak infectiousness. To assess the impact of test sensitivity we also compared this to a 95% sensitivity version of the same model.

**Figure 1b.**
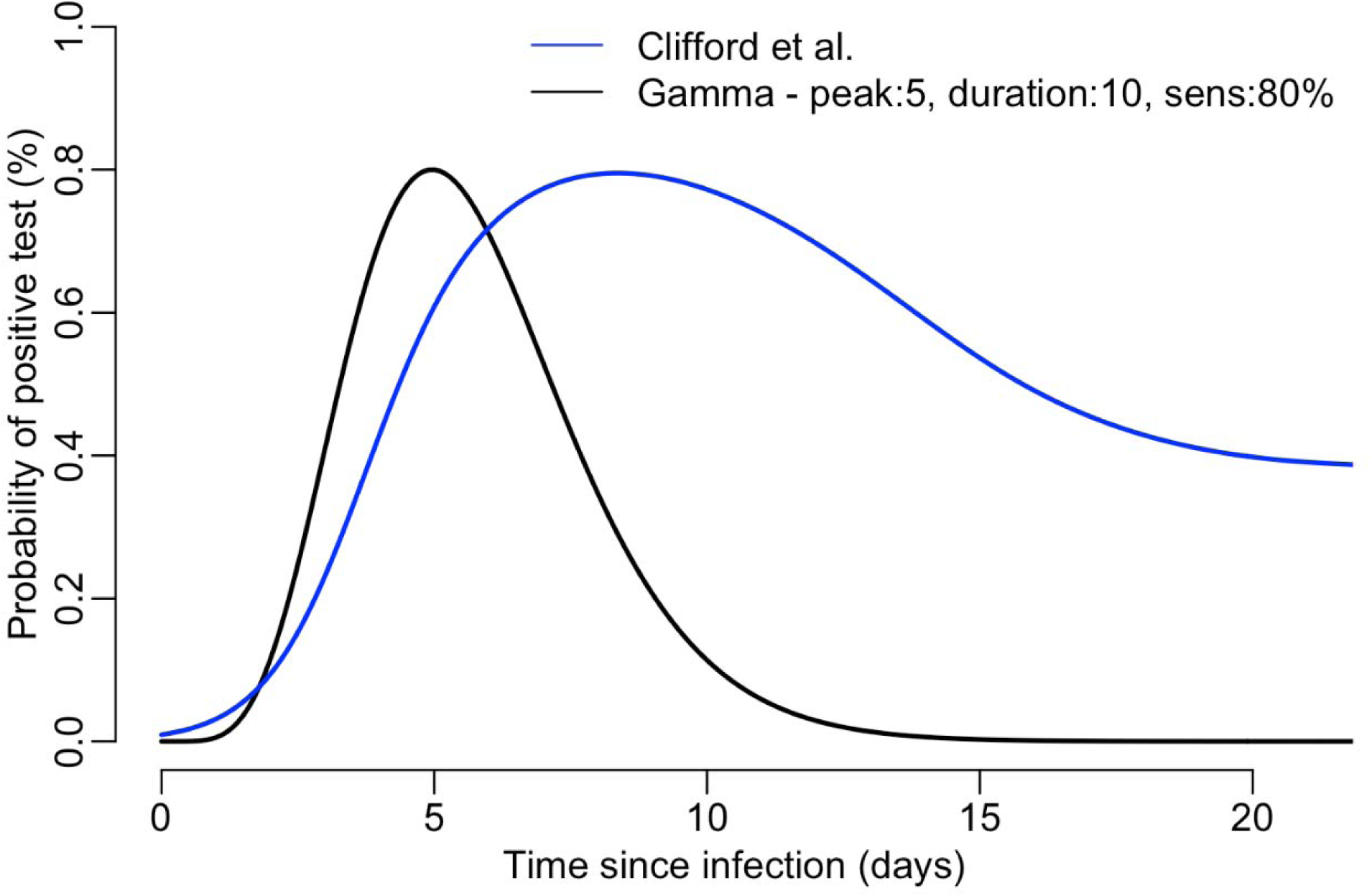
Models of the probability of a positive test for SARS-CoV-2 relative to time since infection: a distribution estimating positivity by RT-PCR adopted from Clifford et al. [44] and the Gamma infectiousness function as an approximation for an antigen test when test positivity tracks infectiousness with a maximum sensitivity of 80%.

We then constructed a model capturing these components to assess the impacts of testing, symptom monitoring, and quarantine. Infections resulting in travel-related risk could occur before or during a trip and we use the infectiousness described above, *I(t)*, at time *t* relative to travel based on infection at time *τ*, relative to travel (prior to travel is negative):

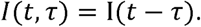

Here, we assessed two transmission windows: Days 0-1 for risk during travel to include potential risk in transit prior to and after airline travel and Days 0-28 for risk after travel.

Symptom monitoring was assessed as a method to detect and isolate infected individuals and therefore prevent transmission after symptom onset. As described above, we assumed that a proportion of infected individuals develop symptoms (*σ*_0_) and develop symptoms at rate *σ(t)* as defined by the incubation period (described above). The onset of symptoms was assumed to lead to isolation until recovery, resulting in a residual in transmission risk over the transmission window:

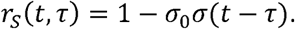

Transmission at a time *t* can also be mitigated through quarantine. We estimated the impact of quarantine as a reduction in risk of a magnitude equal to the adherence ∝_Q_ (1 = 100%) during a quarantine of duration *T*_Q_ starting at the time of arrival *t*_0_ with residual transmission risk:

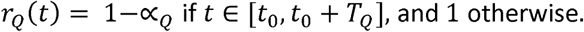

Transmission can also be mitigated through test-based detection followed by isolation. For the purposes of the model, we assumed that test results were immediately available and a positive test immediately led to isolation until recovery. Test positivity for each test (described above) was characterized *ρ*(*t*) and the corresponding residual in transmission associated with each test *k* at test time *t*_*k*_ is:

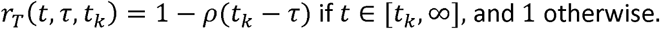

For a set of tests, *K*, the residual risk is the product:

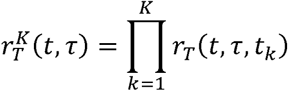

Here, we assessed two transmission windows: Days 0-1 for risk during travel to include potential risk in transit prior to and after airline travel and Days 0-28 for risk after travel. The total transmission risk between times *t*_1_ and *t*_2_ for individuals infected at time *τ* is:

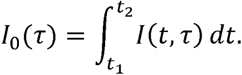

The transmission risk for individuals with protocols including symptom monitoring, quarantine, and testing is:

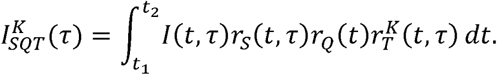

For exposure windows in which a unique time of exposure is unknown, we define the risk of exposure as uniformly distributed over a window defined by *τ*_1_ and *τ*_2_:

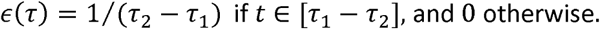

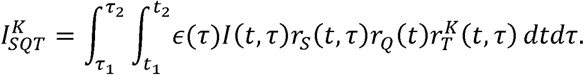

Finally, we calculate the proportional reduction in transmission risk as: 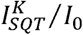

## Results

### Reducing transmission risk after a specific known exposure

Before looking at exposure over a range of times, we first assessed the impact of symptom monitoring, quarantine, and testing when the time of infection was known (for example, a brief high-risk contact). Isolating infected individuals at the time of symptom onset, without testing or quarantine, resulted in a reduction in transmission risk of 36-52% (minimum to maximum accounting for differences in infectiousness over time between models). Quarantine alone led to higher reductions in transmission risk, from 39-75% with 7 days to 90-100% with 14 days. Isolating individuals based on a single positive test result alone produced a 0-77% reduction in transmission, depending on the day of the test relative to the infectious period and the time-specific test sensitivity (Fig. 2). Testing earlier in infection was less effective at detecting infections; later testing means that while the test was more likely to be positive, the infectious period may begin prior to the test, leading to a smaller reduction in risk. Combining symptom monitoring or quarantine with testing provided added benefit, leading to increased risk reduction, especially with a test at day 4-5 post-exposure with symptom monitoring (52-84% reduction) or a test at day 5-6 with a 7-day quarantine (76-99% reduction). A 7-day quarantine with symptom monitoring and a test at day 5-6 further increased the lower bound of likely risk reduction to 91-99%.

**Figure 2.**
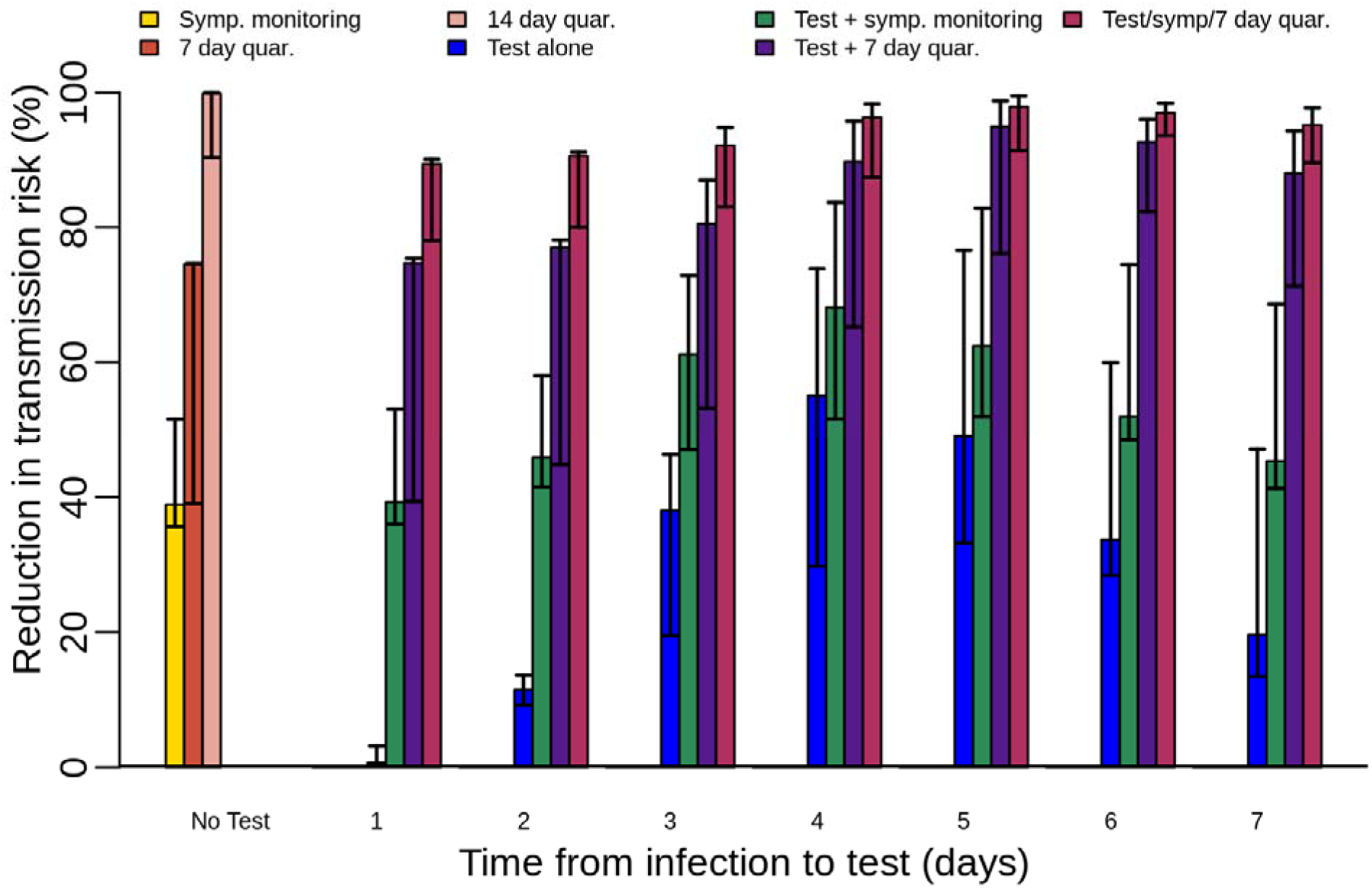
Reductions in total average SARS-CoV-2 transmission risk after infection at a known high-risk exposure time (Day 0), stratified by method of risk reduction and day of test. Symptom monitoring is assumed to be ongoing regardless of the test date when implemented and either symptom onset or a positive test result is assumed to result in immediate isolation until the individual is no longer infectious. The bars represent the median estimate and the error bars show the range (minimum and maximum).

### Transmission risk during travel

To assess approaches for reducing risk of transmission while traveling, we assumed that exposure may have occurred at any time in the 7 days prior to arrival at a destination. Isolating individuals at the time of symptom onset prior to or during travel resulted in a 26-30% reduction in risk (Fig. 3A). Testing resulted in the greatest reduction of risk when the specimen was collected closest to the time of travel. Testing 3 days prior to travel resulted in a 5-9% reduction in transmission risk compared to a 37-61% reduction with testing on the day of travel. This was also true for testing combined with symptom monitoring, which had higher overall reductions.

**Figure 3a.**
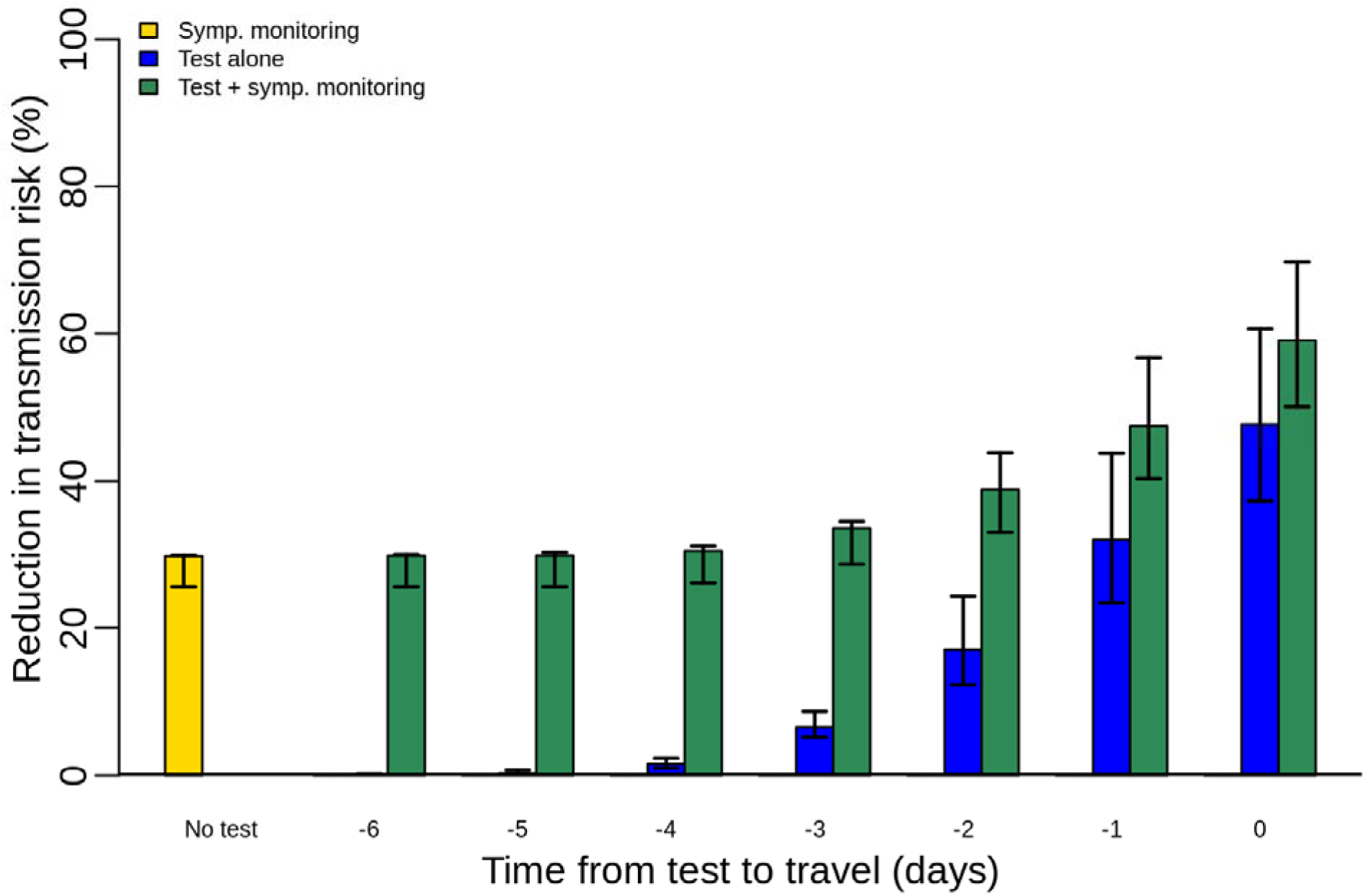
Reductions in SARS-CoV-2 transmission risk during a 1-day trip assuming a 7-day exposure window prior to travel, stratified by method of risk reduction. Individuals developing symptoms are assumed to be isolated and therefore do not travel.

We assessed the impact of test sensitivity relative to timing by comparing the antigen-type test model to the same model with higher sensitivity. With the same time-specific pattern but different sensitivity (80% vs. 95%, Fig. 3B), the higher sensitivity test gives a higher reduction in transmission risk if used at the same time. However, the importance of sensitivity is intertwined with timing. The lower sensitivity test was as effective or more effective than a higher sensitivity test if it was performed closer to the time of travel. For example, the test with 80% sensitivity performed 1 day prior to departure was 36-44% effective at reducing transmission risk during travel, while the test with 95% sensitivity performed 3 days prior to departure was 8-10% effective.

**Figure 3b.**
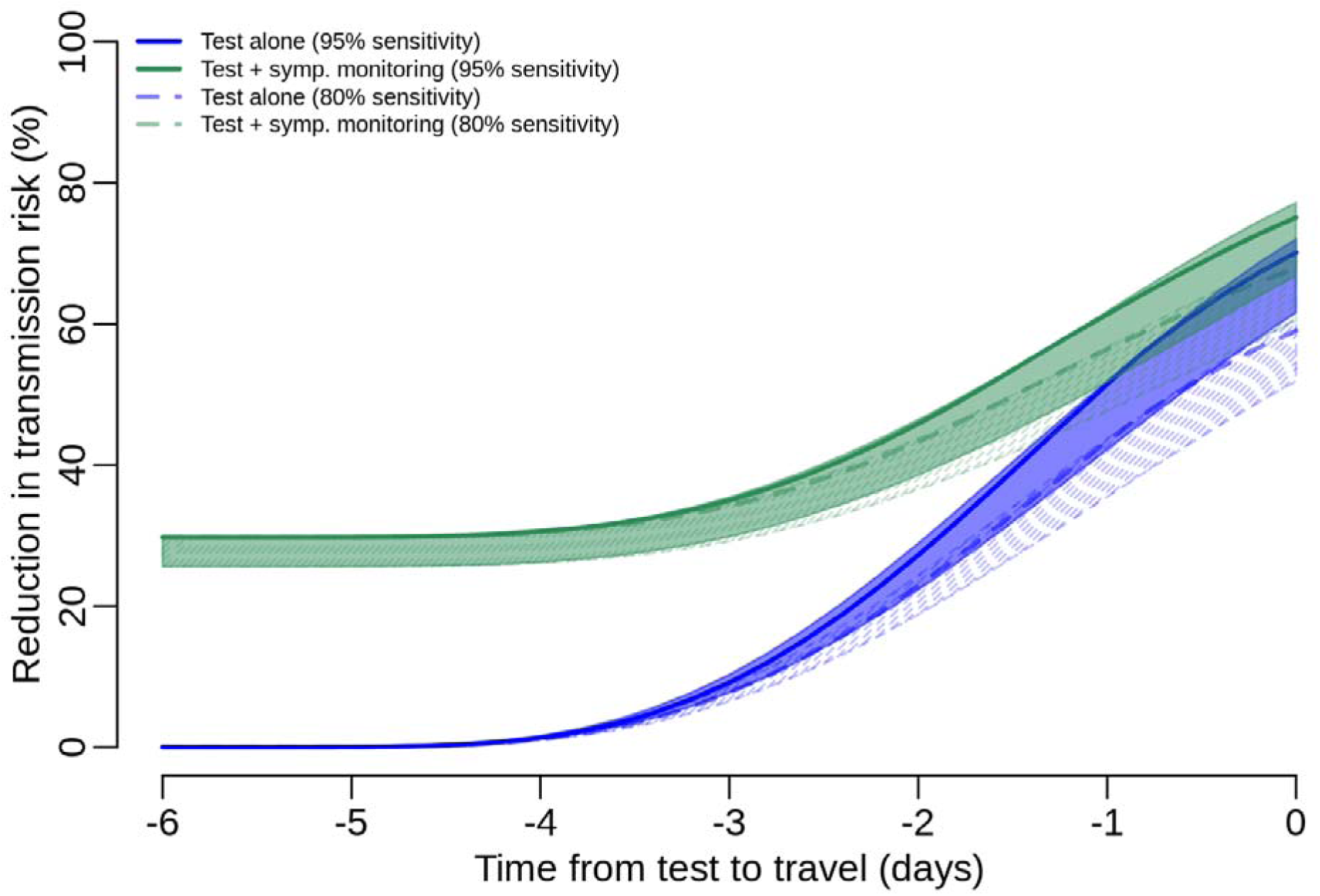
Reductions in SARS-CoV-2 transmission risk during a 1-day trip assuming a 7-day exposure window prior to travel comparing the Gamma function version of the assays with 80% and 95% sensitivity. Ranges indicate uncertainty from the different infectiousness models.

Compared to the effects with a known infection time (Fig. 2), the mitigation measures are generally more effective with a 7-day exposure window (Fig. 3A). This trend continues with longer potential exposure periods prior to travel (e.g., 2 weeks or more). Individuals infected 7 days or more prior to travel may be infectious while traveling, but on average they are less infectious than those infected closer to the time of travel.

### Transmission risk after travel

We then considered measures to reduce the risk of SARS-CoV-2 introduction to the destination location from travelers, i.e., transmission risk after traveling (Fig. 4). Assuming infection occurs at an unknown time within a 7-day exposure period prior to arrival, a single test on its own was most effective when performed 1- or 2-days post-arrival (29-44% and 29-45% reduction in transmission risk, respectively). This reduction in introduction risk was higher than reductions generated by testing prior to travel; a test 1 day prior to arrival provided a 17-30% reduction in risk and a test 3 days prior to arrival provided an 8-21% reduction (not shown). Tests prior to travel do not detect travelers infected while traveling and were less likely to detect travelers infected close to the time of travel. These travelers are those who are most likely to experience their entire infectious period in the destination location, and therefore, pose the greatest introduction risk.

**Figure 4.**
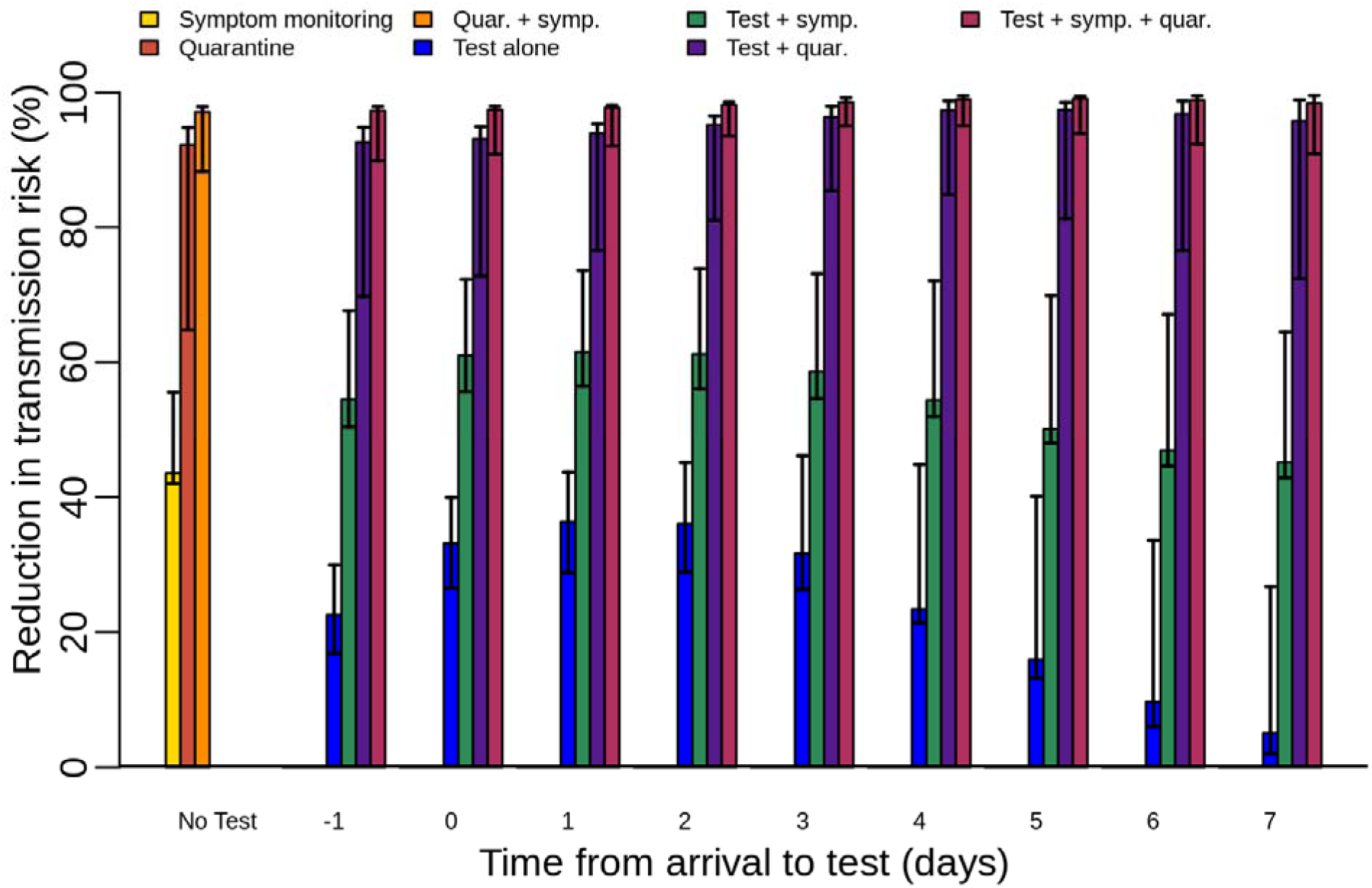
Reductions in SARS-CoV-2 transmission risk from infected travelers post-arrival assuming a 7-day exposure window prior to arrival, stratified by day of test and symptom monitoring, with and without a 7-day quarantine. Symptom monitoring is assumed to be ongoing before, during, and after travel and either symptom onset or a positive test result is assumed to result in immediate isolation until the individual is no longer infectious.

Although a pre-travel test was less effective on its own than a post-travel test, the combination of pre-travel and post-travel tests provided additional risk reduction. A pre-travel test was most effective at reducing transmission risk after travel when performed close to the time of travel (as described above for risk during travel). In the absence of post-arrival quarantine, a second test post-travel was optimal 2-3 after arrival. The pre-travel test was likely to detect individuals who were infectious upon arrival and the later test was likely to detect those who became infectious after arrival. Combined, these tests can reduce introduction risk by 40-66%. A similar effect can be attained by testing immediately upon arrival and again 3-4 days post-arrival, which reduced introduction risk by 45-70%.

Symptom monitoring and isolation before, during, and after travel, with no other measures in place, reduced introduction risk by 42-56% and was more effective when combined with testing (Fig. 4). For example, a test 1-day post-arrival combined with symptom monitoring before, during, and after travel reduced introduction risk by 56-74%. However, quarantine for 7 days or more on its own was more effective than testing combined with symptom monitoring, regardless of when the test occurred. A 14-day quarantine reduced transmission risk by 96-100%, a 10-day quarantine by 84-100%, and a 7-day quarantine by 65-95% (Fig. 5). Testing and symptom monitoring further enhanced the effectiveness of quarantine. A single test conducted 3-4 days after arrival with symptom monitoring and a 7-day quarantine reduced introduction risk by 95-99% (Fig. 4). The day 3-4 window is optimal because it balances the reduced risk while in quarantine, with higher sensitivity for detecting individuals who may remain infectious at the end of the quarantine period.

**Figure 5.**
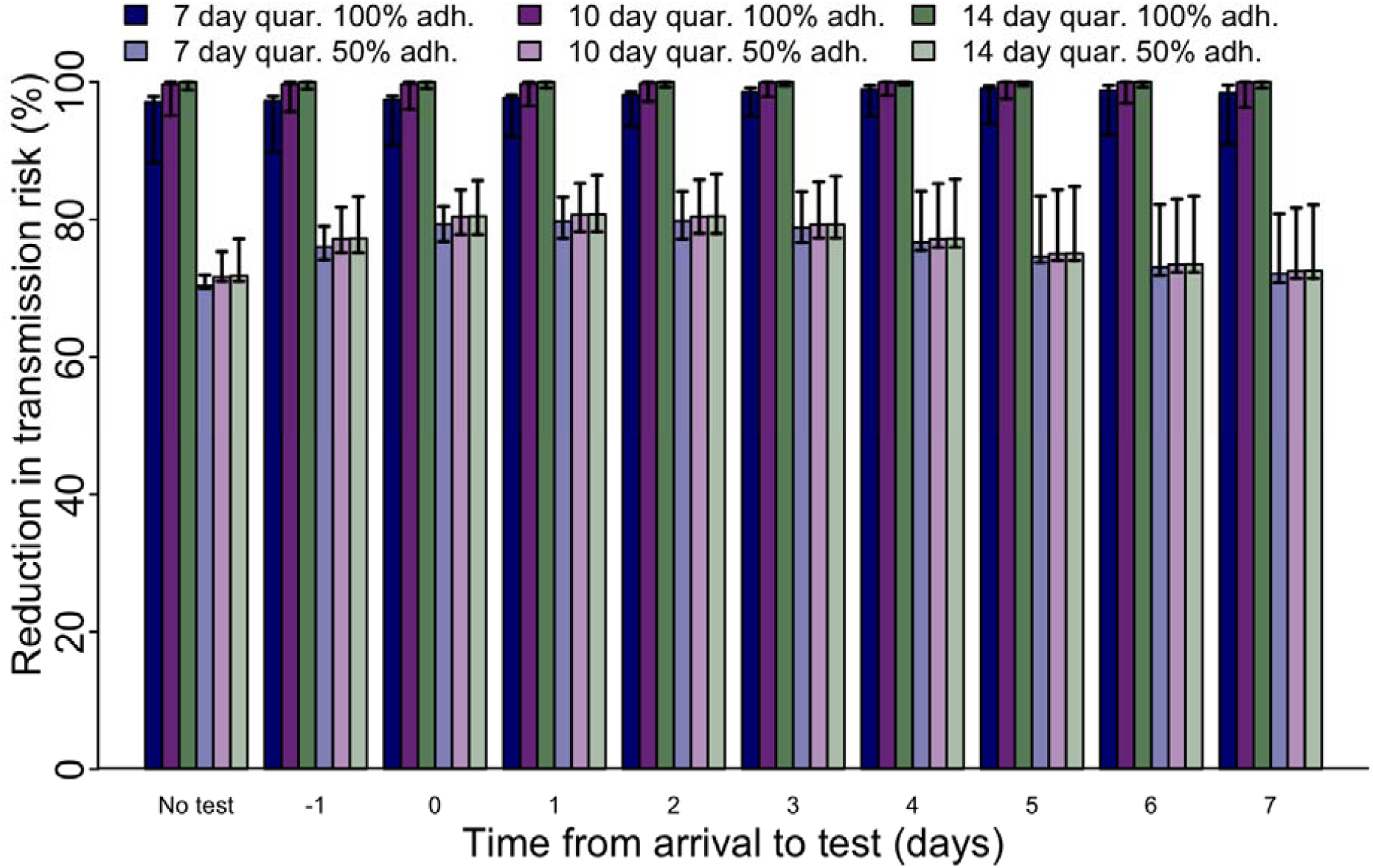
Reductions in transmission risk post-arrival assuming a 7-day exposure window prior to arrival and symptom monitoring, stratified by quarantine length, quarantine adherence, and day of test.

A 7-day quarantine in conjunction with symptom monitoring and testing had similar effectiveness to a 10-day or 14-day quarantine on its own. Comparing quarantine with imperfect adherence (50%), we found that with symptom monitoring and no test, a 7-day quarantine (70-72%) was likely to be almost as effective as a 14-day quarantine (71-77%; Fig. 5). Combined with a test within 0-3 days after arrival and symptom monitoring, a 7-day quarantine with 50% adherence was estimated to be more effective (77-84%) than a 14-day quarantine with 50% adherence and no test (71-77%) and as effective as a 14-day quarantine with a test (77-87%).

## Discussion

Control of SARS-CoV-2 is contingent upon multiple layered mitigation measures. Reducing the risk of transmission associated with travel is critical to reducing the impact related to importations on local health and healthcare systems. This is important when transmission at the destination is low and an introduction could spur additional outbreaks, but also when transmission is already high and health systems may be strained. Reducing risks associated with air travel could pave the way to air industry recovery, as well as offer relief to national economies and reduce social distress [56]. Efforts to control transmission before and after travel rely on individual mitigation measures such as mask use and social distancing before, during, and after travel, but additional control measures, such as testing and quarantine, have also been used by some countries. The fifth meeting of the International Health Regulations Emergency Committee convened by WHO regarding the COVID-19 pandemic stated that for health measures related to international travel, countries should regularly reappraise measures applied to international travel and ensure those measures (including targeted use of diagnostics and quarantine) are risk- and evidence-based [57].

Here, we used a mathematical model to assess the relative impact of three mitigation measures to reduce transmission risk from infected travelers: symptom monitoring, testing, and quarantine. We assessed combinations of these mitigation measures with different estimates of the infectious period, different estimates of test-positivity relative to time of infection, and different assumptions about infection timing and test sensitivity. We frame these results as proportional reductions in transmission risk from infected travelers during or after travel to consider the importance of optimizing mitigation measures to address peak infectiousness (Fig. 1a). On its own, quarantine was the most effective of the three strategies, with a 14-day quarantine almost eliminating risk and a 7-day quarantine being more effective than any single other measure. However, these measures can be more effective when used together. Symptom monitoring is relatively easy and further increases the effect of a 7-day quarantine to 88-97% (with a 7-day exposure window prior to arrival; Fig. 4).

Testing also provides added benefit but is contingent on the timing and quality of the test. Testing prior to travel reduces transmission risk both while traveling and after travel if testing is done close to the time of travel. Testing closer to the time of travel is more likely to detect individuals who are infectious while traveling and immediately afterwards but can still miss infected travelers who are in their latent period, as they may not have enough viral shedding to be detected. While testing immediately prior to travel can substantially reduce risk, it poses additional logistical challenges: results must be reliably available prior to travel and protocols would be needed to effectively isolate individuals who test positive. On the other hand, testing more than 3 days before travel provides little benefit beyond what symptom monitoring can provide, because individuals who test positive at that point contribute less to transmission risk later, including both during and after travel. Because of the value of testing close to the time of travel, a lower sensitivity test with faster results can be more effective despite decreased sensitivity. This finding is consistent with modeling work by Larremore et al. showing that limitations of reduced sensitivity can be overcome by more frequent testing that can still identify infections in time to reduce transmission, in this case closer to the time of travel [58]. This conclusion draws attention to the importance of turnaround times to allow for corresponding decision-making, not just the sensitivity of the test. While test and setting-specific test turnaround times are critical to planning, they are highly varied and were not included here. These results should be considered in that context. For example, short turn-around time is very important for pre-travel testing but less critical for post-travel testing at day 1 or 2 when individuals are expected to remain in quarantine for 7 days or more.

In the absence of quarantine or with low adherence to quarantine, post-arrival testing is likely most effective 1-2 days after arrival, balancing early detection with optimal sensitivity for travelers in their latent period while traveling. With high-adherence quarantine or potential exposure closer to the time of travel (for example, while traveling), optimal post-arrival test timing is later, 3 or more days after travel. This corresponds to improved sensitivity for detecting individuals who may be infected close to the time of arrival and are most likely to be infectious at the end of the quarantine. With exposure up to 7 days prior to travel, we found that optimal test timing was on days 0-2 after arrival with symptom monitoring and no quarantine, days 3-4 with symptom monitoring and quarantine with 100% adherence, and days 0-3 with symptom monitoring and quarantine with 50% adherence. Beyond days 5-8 post-infection, the sensitivity for detecting infections in the models considered here begins to decrease (Fig. 1b). Even with quarantine measures in place, tests on or after arrival may have additional roles if quarantine adherence is imperfect or to assist in contact tracing when other travelers are potentially infected. Waiting to test several days after arrival improves the chance of detecting an individual who will be infectious at the end of the quarantine but does not optimize early detection of other infections among travelers.

These results are generally consistent with other analyses of risk associated with travel. Early in the pandemic, it was apparent that symptom screening at airports or other transit hubs could not stop the spread of SARS-CoV-2 [18]. Using an individual-level simulation framework, Clifford et al. found that more than half of infected travelers would not be detected by exit and entry screening based on temperature measurement, observation for illness, and health declaration [44]. Sufficient detection of infected travelers to avoid uncontrolled importations is largely dependent on a set of assumptions that are inconsistent with COVID-19 epidemiology: asymptomatic transmission being negligible, very high airport symptom screening sensitivity, and a short incubation period. Clifford et al. also assessed combined measures and estimated that an 8-day quarantine period with an RT-PCR test on day 7 would be nearly as effective as a 14-day quarantine on its own. Other recent work highlights the effectiveness of shorter quarantine periods combined with testing for individuals with known exposures [46,59,60]. Across these studies, the specific days for quarantine or testing and the estimated effectiveness varied due to differences in assumptions about the time of exposure, different modeled test characteristics, and differences in parameters for the infectious period. Nonetheless, all indicate the value of shorter quarantine combined with symptom monitoring and testing, a finding that is helpful both in the travel setting and in other settings with exposure risk.

The model used here has some specific limitations. First, the infectious period of SARS-CoV-2 is not well-defined. We considered multiple models of the infectious period generated by multiple approaches to reflect uncertainty around this period, yet these models also have limitations, are not exhaustive, and more detail is needed for more precise estimates. Moreover, each of the infectious period models captures only the average infectious period, so for individual travelers, this could be substantially different. The most effective measures modeled here are close to 100% effective in the model; however, the existence of individual-level variation suggests that none of these approaches would truly be 100% effective. Even with a 14-day quarantine, it is likely that some individuals will be infectious later, or even develop symptoms only at the end of the time period. Nonetheless, the average parameterization gives the expected average effectiveness for larger numbers of infected travelers; this is the scale at which policies may be most useful. Testing options are also highly varied and not well-characterized. The test options considered here are not exhaustive nor precisely characterized. Moreover, test turn-around time can also vary. We did not model test turn-around time; instead we focused on when the test was performed, such that the results could be considered in the context of whatever testing and laboratory resources are available. For example, a test during quarantine should be done sufficiently early so that results are available before the end of quarantine, but that delay varies in different settings. Our framework, however, can be applied with many other options, or with better characterized distributions as these become available.

We also did not consider behavioral aspects of prevention, with the exception of adherence to quarantine. For simplicity, we assumed that quarantine was equivalent to individual-level isolation and that symptomatic individuals or those testing positive are also isolated immediately. However, individuals may quarantine with others. In that case, symptom onset or a positive test for a single individual can indicate exposure for the others during quarantine. Without symptom onset or a positive test, there may be silent secondary transmission that could result in additional post-quarantine risk. Moreover, travelers may have little incentive to consistently adhere to these measures, and notification or enforcement of them also would require substantial effort and resources. Some travelers could attribute symptoms to other etiologies, such as an exacerbation of a pre-existing condition or travel fatigue. While adherence to all measures may be lower in practice than considered here, the relative effectiveness of measures still provides a useful guide. Moreover, the effectiveness of shorter quarantines, especially when combined with symptom monitoring and testing, may be enhanced because a shorter quarantine is less onerous and may drive better adherence [61].

Finally, we focused on comparing the effectiveness of intervention measures for infected travelers. This is not an analysis of the conditions in which these measures should be implemented, nor of the specific logistical and policy challenges that arise in different situations. Quarantine of all travelers can be an effective prevention measure but could also result in the restricted movement of many travelers who are not infected and, therefore, pose no risk. When the absolute risk of infection in travelers is low and the number of travelers is high, quarantine of travelers without symptoms would predominantly result in the quarantine of uninfected people. Testing is helpful in part because it can reduce the length of quarantine needed for optimal prevention. However, testing can also result in false negatives (missed cases that are released from quarantine when still infectious) or false positives (individuals who test positive but are not actually infected). The impact of false positives can be partly mitigated by confirmatory testing. It is also possible that some recently recovered individuals will test positive but no longer be infectious (e.g., by RT-PCR which can detect SARS-CoV-2 RNA after the infectious period has ended). Additional testing or assessment of cycle threshold values may help reduce the impact on these individuals [62]. It is important that authorities also carefully consider prioritization of testing resources in the context of other public health needs in resource-limited situations.

A multi-layered approached is needed to control SARS-CoV-2 transmission associated with travel. Infection prevention measures (e.g., social distancing, mask use, hand hygiene, enhanced cleaning and disinfection) are expected to reduce risk before, during, and after travel. Symptom monitoring, quarantine, and testing can all complement those measures to further reduce risk. Pre-departure SARS-CoV-2 testing can supplement symptom monitoring to identify potentially infectious travelers who do not have symptoms, and therefore, offers an opportunity to further reduce transmission risk during and after travel. Post-arrival SARS-CoV-2 testing can identify asymptomatic or pre-symptomatic infected travelers, including some who may have tested negative prior to departure, if prior testing took place. Post-arrival testing is likely effective at days 1-2 without quarantine, but more effective later, at days 3-4, if effective quarantine is in place. A 14-day quarantine is effective on its own but combined with testing and symptom monitoring (with isolation of those who develop symptoms or test positive), quarantine can be shortened and still be effective. These findings can inform policies for travel until safe and effective vaccines become widely available.

## Data Availability

Only publicly available data informed this research.

